# A photoplethysmography-based aging clock reveals genetic determinants of arterial aging

**DOI:** 10.1101/2025.11.15.25340310

**Authors:** Lingling Xu, Lanyue Zhang, Anushree Ray, Murad Omarov, Rainer Malik, Nathalie Beaufort, Martin Dichgans, Nicola Luigi Bragazzi, Marios K. Georgakis

**Author notes:** **Correspondence**: Marios K. Georgakis, MD, PhD, Institute for Stroke and Dementia Research (ISD), Ludwig-Maximilians-University (LMU) Hospital, Feodor-Lynen-Str. 17, 81377 Munich, Germany.

## Abstract

Arterial aging, marked by progressive vascular stiffening, is a contributor to cardiovascular disease. Photoplethysmography (PPG) waveforms offer an easily accessible signal of arterial function, enabling scalable assessment of arterial age at the population level. Here, we present a multimodal deep learning framework integrating raw PPG waveforms with hemodynamic features from UK Biobank participants to construct an arterial aging clock. From this model, we derived *ArtAgeGap*, an age-independent residual biomarker of arterial aging, which correlated with established measures of vascular stiffening, such as pulse pressure (*r*=0.60). *ArtAgeGap* was significantly elevated in individuals with a history of cardiovascular disease, and associated with diabetes (+0.93 years, 95%CI: 0.64-1.21) and hyperlipidemia (+0.49, [0.36-0.61]). In 96,615 participants followed for a median of 13 years, higher *ArtAgeGap* values predicted incident hypertension, major adverse cardiovascular events, and cardiovascular and all-cause mortality (hazard ratios per +1 year: 1.01-1.06), on top of chronological age. A genome-wide association study in 114,098 individuals identified 60 independent loci associated with *ArtAgeGap*, including variants previously associated with blood pressure (e.g. *NPR3*), arterial stiffness (*CLCN6*), aortic diameter (*SLC24A3*), and atherosclerosis (*HDAC*9), as well as 34 novel loci enriched for arterial tissue expression. Integrative analyses with transcriptomic data from human arteries prioritized 28 genes, such as *RSG19* and *ULK4*, whose genetically proxied expression was associated with *ArtAgeGap*. Single-cell transcriptomic data from arterial tissue revealed strong enrichment of these genes in fibroblasts, implicating fibrotic remodeling mechanisms in arterial aging. Rare variant burden testing further implicated damaging variants in *COL21A1*, *LMNA*, *TP53BP2*, *RXRB*, and *FLOT2*, also converging to mechanisms related to extracellular matrix organization and fibrosis regulation. Lastly, Mendelian randomization analysis identified *ArtAgeGap* as an intermediate biomarker in-between vascular risk factors and outcomes, with central adiposity, higher blood pressure, and type 2 diabetes increasing *ArtAgeGap*, and higher *ArtAgeGap* elevating the risks of coronary heart disease and stroke. Together, these findings establish *ArtAgeGap* as a scalable PPG-based biomarker of arterial aging and provide mechanistic insights into potential therapeutic strategies to mitigate arterial aging.

## Main

Cardiovascular disease remains a leading cause of death and disability globally^1,2^. Age is the dominant risk factor for the arterial pathologies that drive cardiovascular disease^3^. Aging is associated with profound functional and structural changes in the human vasculature, most notably progressive stiffening of the large arteries^4,5^. Arterial stiffness elevates systolic blood pressure and reduces the arteries’ capacity to buffer the pulsatility generated by the heart^6^. The resulting loss of cushioning function allows excessive pulsatile energy to penetrate into the microvasculature of end organs such as the kidney, retina, and brain, promoting target organ damage. While acknowledged as a high-priority therapeutic target for lowering the burden of cardiovascular disease, there are no effective interventions to ameliorate arterial aging and stiffness^7^.

Arterial stiffness is most commonly assessed using pulse wave velocity (PWV), a biomarker robustly associated with future cardiovascular and cerebrovascular events^8,9^. Carotid-femoral PWV (cfPWV) is the most validated and widely accepted metric^10^ and is typically quantified with non-invasive tonometry approaches. However, cfPWV measurement requires specialized equipment and trained operators, limiting its feasibility for large-scale application^11^. This creates an unmet need for scalable methods to assess arterial aging that would enable genetic and multi-omics studies of underlying mechanisms and help identify therapeutic targets. Photoplethysmography (PPG), a simple, low-cost, and widely accessible optical technique, offers a promising alternative. By detecting blood volume changes in the microvasculature, PPG enables derivation of surrogate markers of arterial stiffness, including PPG-based estimates of PWV, and supports continuous vascular monitoring in both clinical and consumer settings^12^.

Recent advances in aging clocks derived from imaging and molecular biomarkers have highlighted the potential of data-driven approaches to quantify biological aging. Pulse waveforms, the foundation PWV measurement, are rich in hemodynamic information and may encode deeper insights into arterial aging when analyzed directly using deep learning. However, early studies applying convolutional neural networks (CNNs) to pulse waveform data were constrained by relatively small sample sizes. These studies either demonstrated only modest accuracy in predicting chronological age without downstream epidemiological validation, or they focused on classifying cardiovascular disease states without achieving a fine-grained, year-level estimation of “arterial age.”^13,14,15^. A recent study using continuous PPG wearable data demonstrated high accuracy in predicting chronological age, but lacked access to additional omics data to explore mechanisms underlying arterial aging^16^.

In this study, we developed a multimodal deep learning model that integrates PPG-derived pulse waveform data with complementary hemodynamic variables to predict arterial age. We derived a novel chronological age-independent biomarker, *ArtAgeGap,* and validated it as an indicator of arterial aging by correlating it with established vascular traits and longitudinal cardiovascular and cerebrovascular outcomes. To uncover underlying biological mechanisms, we performed extensive genetic analyses, including genome-wide association studies (GWAS) of common variants, as well as gene burden tests based on rare exonic variants derived from whole exome sequencing (WES) data. We followed-up on promising signals with transcriptome-wide association studies (TWAS) and colocalization, and explorations of gene expression patterns in bulk and single-cell RNA sequencing data from human arteries. We also investigated the associations from vascular risk factors to *ArtAgeGap*, and *ArtAgeGap* to cardiovascular adverse outcomes with genetic instruments in Mendelian Randomization (MR) analyses. Based on these analyses, we highlight promising therapeutic targets supported by converging evidence from genetic and omics studies for future experimental exploration.

## Results

### Study overview

Our study design is summarized in **Figure 1a**. Utilizing data from PPG pulse waveforms, waveform-derived characteristics, Blood Pressure (BP), pulse rate, and height as input features, we trained a model to predict chronological age in data from 14,315 individual participant visits in the UK Biobank (**Figure 1b**). To integrate multi-modal data, we designed a dual-branch neural network architecture composed of a CNN for processing image representations of PPG waveforms and a multilayer perceptron (MLP) for structured tabular input. From the trained model, we derived a novel biomarker termed *ArtAgeGap*, defined as the age-adjusted difference between the model-predicted arterial age and actual chronological age. This metric quantifies the degree of arterial aging acceleration or deceleration relative to individuals of the same chronological age. We assessed the interpretability and validity of *ArtAgeGap* as a marker of arterial aging by correlating it with established PPG waveform features of arterial stiffness and cardiovascular risk factors. Furthermore, we evaluated its prognostic utility by assessing associations with future cardiovascular events over a longitudinal mean follow-up period of 13 years. To investigate the genetic architecture of arterial aging, we conducted a GWAS using genotyping data and gene-based rare variant burden testing using WES data in 114,098 and 110,326 individuals of primarily European ancestry analyses. To prioritize promising anti-aging therapeutic targets, we complemented these analyses with TWAS and explorations of age-related patterns of gene expression in bulk and single-cell RNA sequencing in human arterial tissue. Additionally, we also conducted MR to explore the associations between vascular risk factors and *ArtAgeGap*, *ArtAgeGap* and cardiovascular adverse outcomes with genetic instruments. Together, we triangulated convergent lines of multilayered genetic evidence for detecting causal genes driving arterial aging.

**Figure 1.**
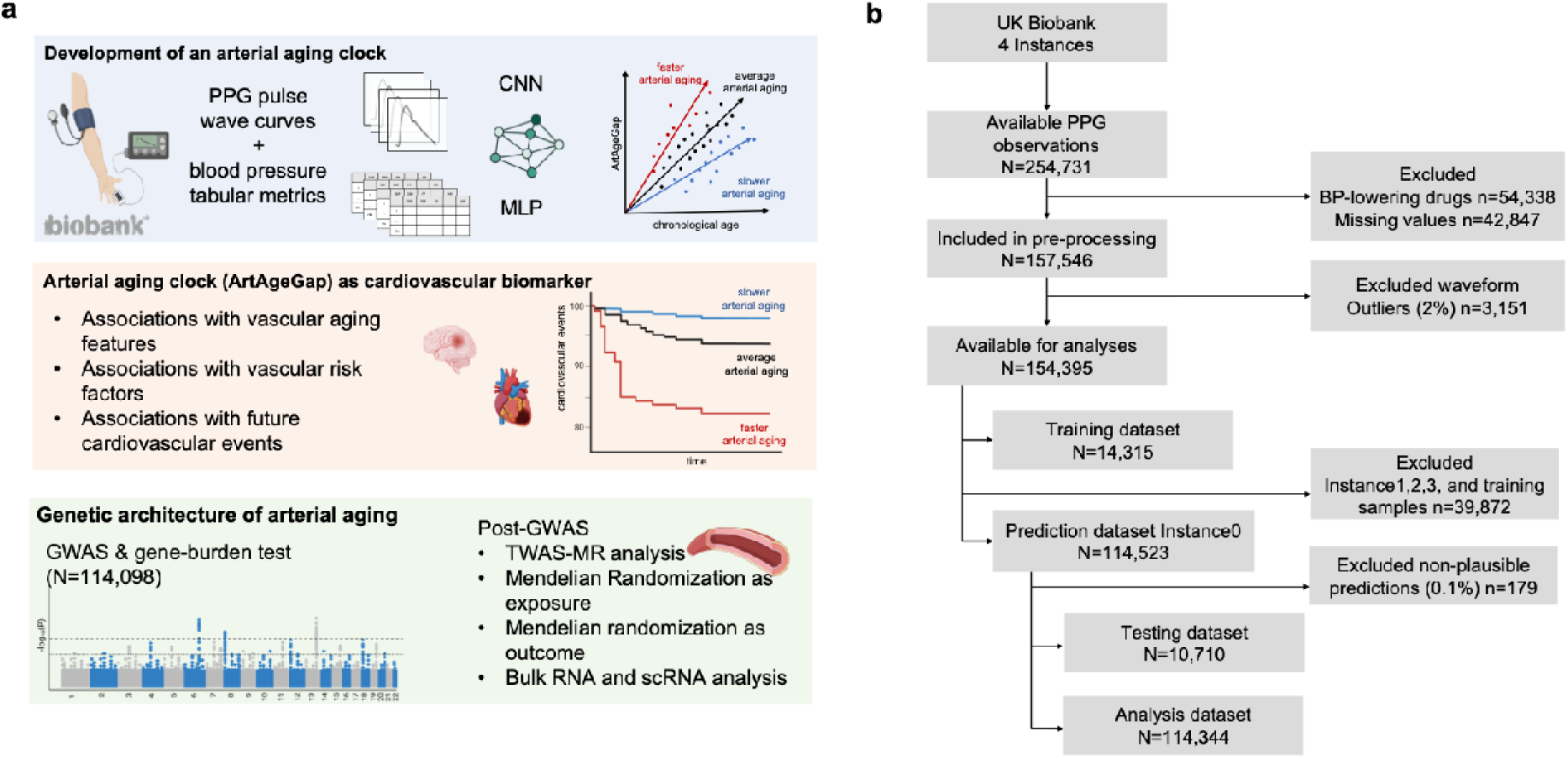
Workflow and flowchart of study participants. (a) Schematic overview of the arterial aging model development and downstream analyses. A deep learning framework combining a convolutional neural network (CNN) and a multi-layer perceptron (MLP) was trained using photoplethysmography (PPG) waveforms, blood pressure, and related physiological features from UK Biobank participants to predict chronological age. *ArtAgeGap* was defined as the adjusted difference between predicted age and chronological age, serving as a biomarker of arterial aging. We assessed the clinical validity of *ArtAgeGap* through its associations with PPG waveform features, established cardiovascular risk factors, and incident cardiovascular events using survival models. To elucidate the genetic basis of arterial aging, we performed a genome-wide association study (GWAS) (n = 114,098) and rare variant burden testing, followed by post-GWAS analyses transcriptome-wide association study-mendelian randomization (TWAS-MR),pathway enrichment, and bidirectional Mendelian randomization. (b) Flowchart of sample selection. From 254,731 UK Biobank observations across 4 time-visits for 219,453 study participants, we excluded PPG waveforms from participants on antihypertensive medications at the time of assessment (n = 54,338), those with missing covariates (n = 42,847), as well as outliers (n = 3,151), resulting in a refined cohort of 154,395 observations across 4-time visits. Of these, 14,315 observations across all visits were used for model development and 10,710 observations were used for model testing. Following exclusion of outlier predictions, the remaining 114,344 observations from the baseline visit (instance 0) were used for further downstream analyses. Each filtering step and the corresponding final cohort sizes are depicted.

### Model performance and development of *ArtAgeGap*, a biomarker of arterial aging

Our training set included 14,315 observations from 4 visits, corresponding to 14,160 study participants aged 40-74 years (median 57 years, 54.6% female), of whom 16.7% (n=2,384) had a self-reported history of cardiovascular disease (**Supplementary Table S1**). Our testing set included 10,710 observations, all from the baseline visit of 10,710 participants aged 40-70 years (median 55 years), 12.8% (n=1,368) of whom had a history of cardiovascular disease (**Supplementary Table S2**). In the training set (**Supplementary Figure 1**), predicted arterial age showed a strong linear relationship with chronological age (R^2^=0.43, r=0.66), yielding a mean absolute error (MAE) of 5.95 years and a root mean square error (RMSE) of 7.42 years. On the held□out testing set, performance remained robust (R^2^=0.33, r=0.60; MAE=5.61 years; RMSE=7.02 years), confirming that the predictor can generalize well to unseen data (**Figure 2a**). The model performed substantially better than a model including only BP-derived features (systolic blood pressure, diastolic blood pressure, and pulse pressure, R^2^=0.16, r=0.44; MAE=6.44 years; RMSE=7.88 years), highlighting the added value of including the PPG waveforms. As expected, the predicted arterial age was consistently 1-2 years older among individual with self-reported cardiovascular disease across the chronological age spectrum (**Figure 2b, Supplementary Figure 2**).

**Figure 2.**
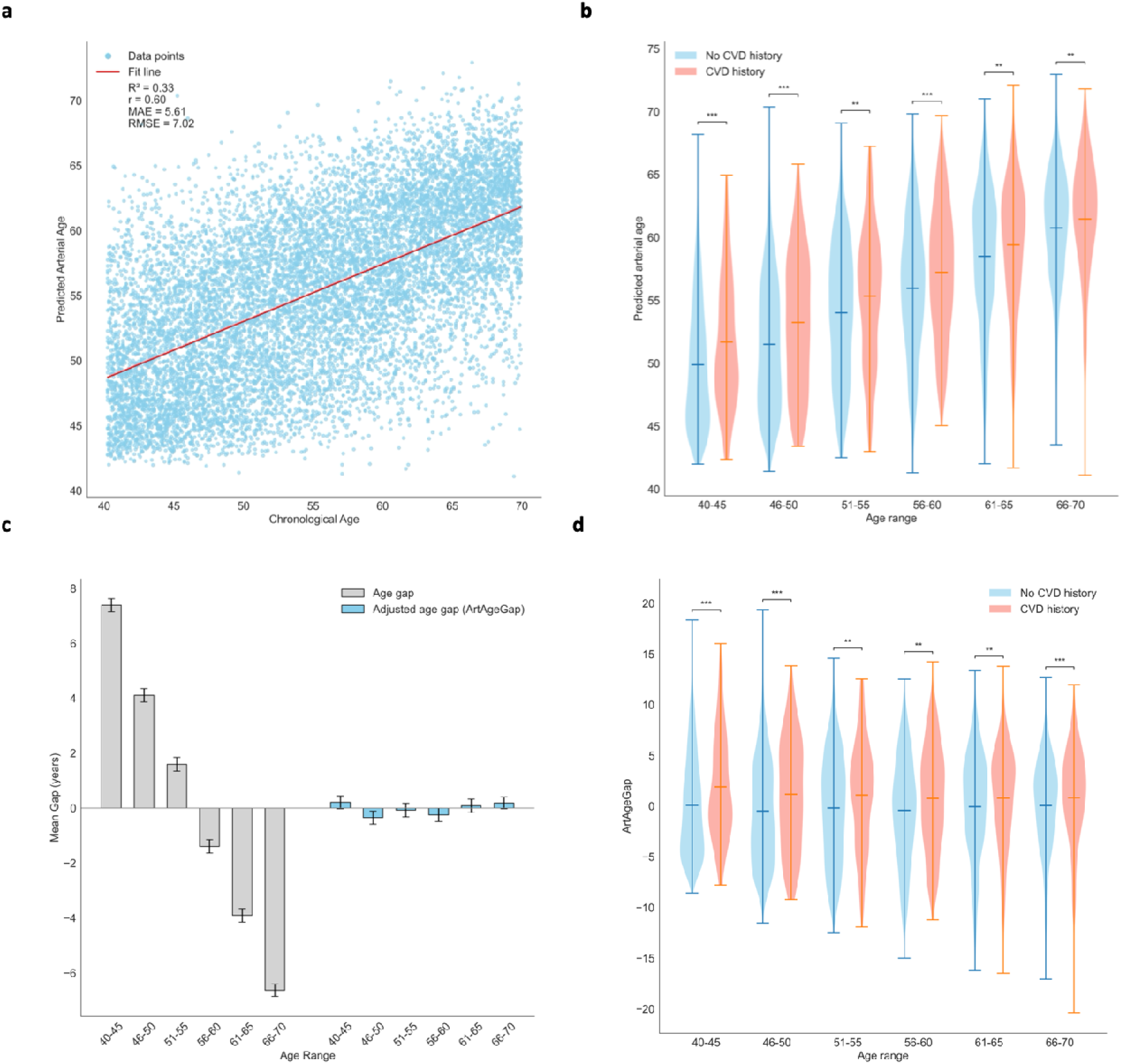
Model performance and age bias correction. (a) Scatter plots of predicted arterial age versus chronological age in the independent testing dataset (n = 10,710), showing model fit with coefficient of determination (R²), mean absolute error (MAE), and root mean square error (RMSE). (b) Violin plots of predicted arterial age across five-year chronological age bins (40-45 to 65-70), stratified by disease history in the testing dataset. Blue violins represent participants without self-reported cardiovascular disease (“No CVD history); orange violins represent those with angina, myocardial infarction, stroke, or hypertension (“CVD history”). Lines indicate bin-wise means. The bar above the violin plots and * shows the significance from Mann-Whitney U test (p<0.05*, p<0.01**, p<0.001***). (c) Raw prediction errors (predicted arterial age - chronological age; gray bars) displayed a systematic age-dependent bias, with younger individuals overestimated and older individuals underestimated. This trend is eliminated after subtracting the fitted quadratic age correction term, yielding the bias-adjusted biomarker *ArtAgeGap* (blue bars centered at zero; bars represent 5%-95% confidence intervals). (d) Violin plots of *ArtAgeGap* across age bins and disease history in the test set. Lines represent mean values within each bin. The bar above the violin plots and * shows the significance from Mann-Whitney U test (p<0.05*, p<0.01**, p<0.001***).

Due to a well-described regression to the mean phenomenon^17^, the age gap generated by subtracting chronological age from our model’s predicted arterial age, exhibited a pronounced negative correlation with chronological age (**Figure 2c**). The predicted arterial age tended to be overestimated in younger participants and underestimated in older ones. To correct for this bias, we applied a quadratic regression model of the unadjusted age gap on chronological age, resulting in coefficients of −0.51 for chronological age, 0.0005 for age squared, and an intercept of 29.79. The resulting age-adjusted arterial age gap (*ArtAgeGap*) was centered around zero across the entire range of chronological ages (Figure 2c). Again, individuals with a history of cardiovascular diseases (CVD) consistently exhibited a positive *ArtAgeGap* of approximately 1-2 years (p <0.01), confirming that CVD correlates with accelerated arterial aging independent of chronological age (**Figure 2d**).

### *ArtAgeGap* is correlated with arterial aging features and cardiovascular risk factors

We next explored, in the remaining UK Biobank dataset with available PPG waveforms in baseline (analysis dataset), if *ArtAgeGap* is correlated with established features of arterial aging (n=114,344, **Figure 3a-b)**. Pulse pressure, a well-established marker of arterial stiffening^5^, showed the strongest positive correlation (ρ=0.60, p <1×10^−4^), followed by systolic blood pressure (ρ=0.42, p <1×10^−4^). Two pulse□wave timing metrics, the time of the shoulder on the waveform (ρ=0.35, p <1×10^−4^) and the time to the primary wave peak (ρ=0.34, p <1×10^−4^), also correlated moderately with *ArtAgeGap*, as was the arterial stiffness index (ρ=0.23, p <1×10^−4^). Conversely, peak□to□peak time (d-b in **Figure 3a**, ρ=-0.25, p <1×10^−4^) was inversely associated with *ArtAgeGap*, whereas diastolic blood pressure did not linearly correlate with *ArtAgeGap* (ρ=-0.004, p=0.23) (**Supplementary Table S3**). These data support that *ArtAgeGap* captures information related to arterial stiffening.

**Figure 3.**
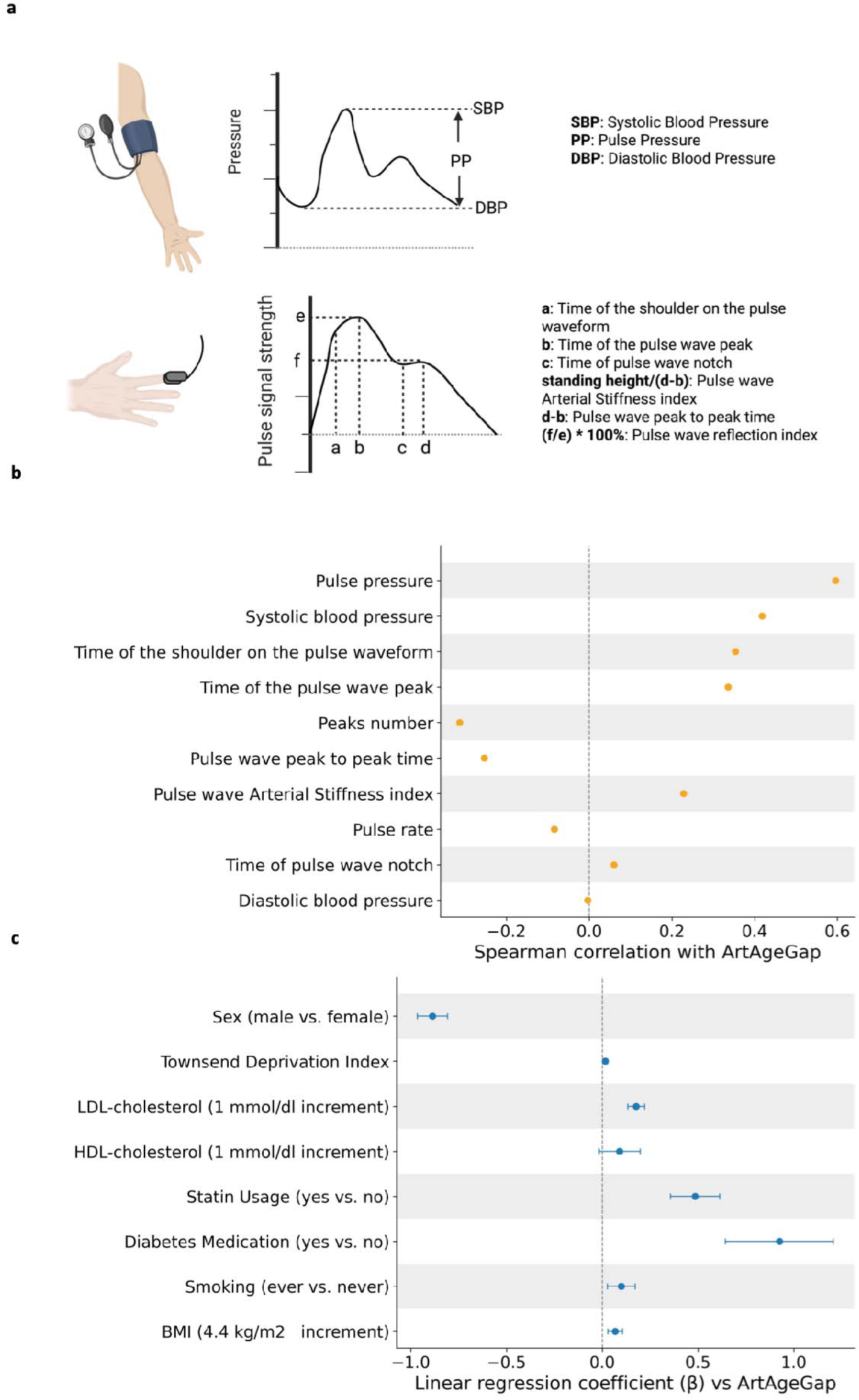
Summary of *ArtAgeGap* Associations with Vascular Features, Cardiovascular Risk, and Future cardiovascular disease (CVD) Events. (a) Schematic illustration of blood pressures and key pulse-wave features extracted from blood pressure examination and photoplethysmography (PPG) signals used in model development and correlation analysis. The figure is generated by Biorender. (b) Forest plot of spearman correlation coefficients (ρ) between *ArtAgeGap* and selected vascular aging phenotypes in the analysis dataset. All marked associations are statistically significant (p<0.05) except Diastolic blood pressure. (c) Forest plot of multivariable linear regression coefficients (β) and 95% confidence intervals for predictors of *ArtAgeGap* (n = 80,278).

In a multivariable linear regression model in the analysis set (n=80,278 with available data for all risk factors), we found significant associations of most established cardiovascular risk factors with accelerated arterial aging (**Figure 3c**). History of diabetes, as captured by prescription of anti-diabetic medications, showed the largest effect being associated with an increase of almost one year in *ArtAgeGap* (β=0.93 years, SE=0.14; p=1.46×10^−10^). Statin use, probably capturing history of hyperlipidemia or atherosclerotic cardiovascular disease (β=0.49 years, SE=0.07; p=1.43×10^−13^), as well as higher LDL-cholesterol at baseline (β per mmol/dl increase=0.18, SE=0.02; p=3.1×10^−15^) were also associated with increases in *ArtAgeGap*. Furthermore, history of smoking (β=0.10 years, SE=0.04; p=0.007) and higher body-mass index (BMI) (β per 4.4 kg/m² increment=0.07 years, SE=0.02; p=5.6×10^−4^) conferred modest increases in *ArtAgeGap* (**Supplementary Table S4**). Notably, despite the inclusion of all covariates, this model of demographic and cardiovascular risk factors accounted for only 1% of the variance in *ArtAgeGap* (R^2^=0.01).

### *ArtAgeGap* is predictive of incident hypertension, adverse cardiovascular events, and mortality

To investigate the prognostic value of *ArtAgeGap*, we analyzed data from 96,615 participants in the analysis dataset, who were followed-up for a median of 12.7 years (IQR 12.4-13.0 years). In Cox proportional hazard models adjusted for chronological age, sex, previous and current smoking, statin usage, diabetes medication, Townsend deprivation index, and BMI, we explored associations from 96,615 participants with all-cause mortality (5,603 events), cardiovascular death (850 events), major adverse cardiovascular events (MACE) (6,337 events), acute myocardial infarction (1,685 events), stroke (1,664 events), unstable angina (503 events), heart failure (1,635 events) and hypertension (17,381 events). On top of chronological age, each year increase in *ArtAgeGap* was associated with 1-6% higher risk of all outcomes (**Figure 4a**). The strongest hazard ratios (HR) were observed for hypertension (HR per year increment 1.06, 95% CI 1.06-1.06) and cardiovascular death (HR per year increment 1.03, 95% CI 1.02-1.05). In a sensitivity analysis in a subsample of participants with repeated BP measurements with the same protocol in follow-up visits of the UK Biobank (n=8,590 participants), we were able to confirm the association with new-onset hypertension (2,597 cases; HR: 1.05, 95%CI: 1.04-1.06). Kaplan-Meier curves showed dose-dependent associations of *ArtAgeGap* with MACE, cardiovascular death, and all-cause mortality (**Figure 4b-d**, **Supplementary Table S5-13**).

**Figure 4:**
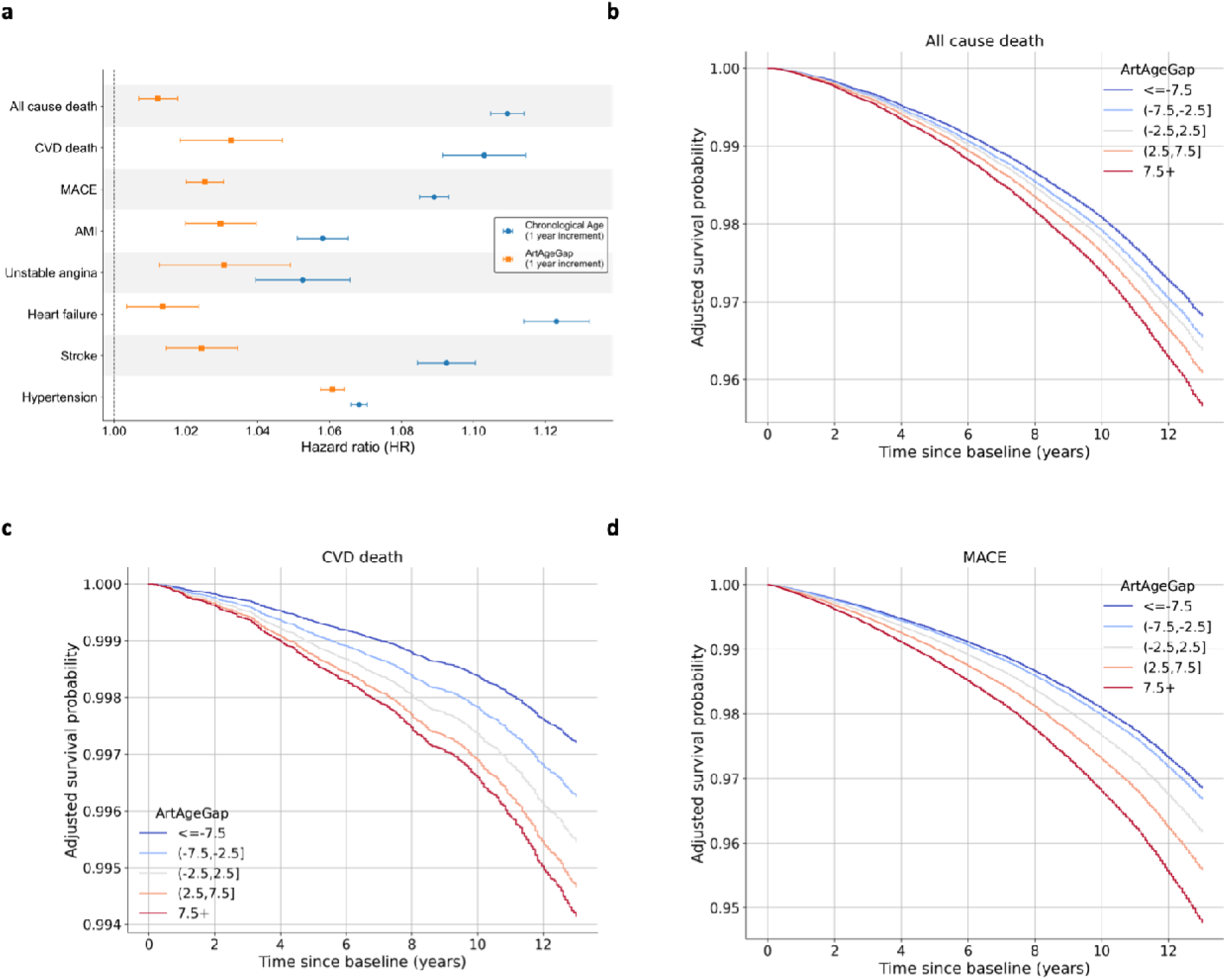
Associations of *ArtAgeGap* with Cardiovascular Outcomes and Mortality. (a) Hazard ratios (HRs) per 1-year increase in *ArtAgeGap* and chronological age from Cox proportional hazards models for multiple endpoints: all-cause death, cardiovascular (CVD) death, major adverse cardiovascular events (MACE), acute myocardial infarction (AMI), stroke subtypes, unstable angina, heart failure and hypertension (n = 96,615). (b-d) Kaplan-Meier survival curves for all-cause death (b), CVD death (c) and MACE (d) by *ArtAgeGap* category (<= -7.5, -7.5 to -2.5, -2.5 to 2.5, 2.5 to 7.5, > 7.5 years), adjusted for chronological age and sex.

### Genetic Architecture of *ArtAgeGap*

To explore biological mechanisms underlying arterial aging, we next conducted a GWAS (**Figure 5a**) of *ArtAgeGap* in participants of the analysis set with available genotyping data (n=114,098). Following filtering for frequency (minimum allele frequency (MAF) >1%) and imputation quality (INFO>0.3), 10,224,488 variants were analyzed. Our GWAS revealed 4,807 genome□wide significant signals (p <5×10^−8^, **Supplementary Table S14**), which collapsed after LD clumping (r² < 0.001 within 10 Mb) into 60 independent loci (**Supplementary Table S15**). The single nucleotide polymorphism (SNP) based population heritability for *ArtAgeGap* was 18%. Significant signals included established variants previously associated with BP (e.g. *NPR3*), arterial stiffness (*CLCN6*), aortic diameter (*SLC24A3*), and atherosclerosis (*HDAC*9). However, 34 of the loci (e.g. *LINC02398* and *ULK4*) were not previously associated with vascular traits, representing potentially novel loci. Across 54 tissue types in the Genotype-Tissue Expression (GTEx) project, the top tissues enriched for the upregulated expression of genes closest to the respective loci were the tibial artery, the aorta, the coronary artery, and the heart atrial appendage (**Supplementary Figure 3)**.

**Figure 5.**
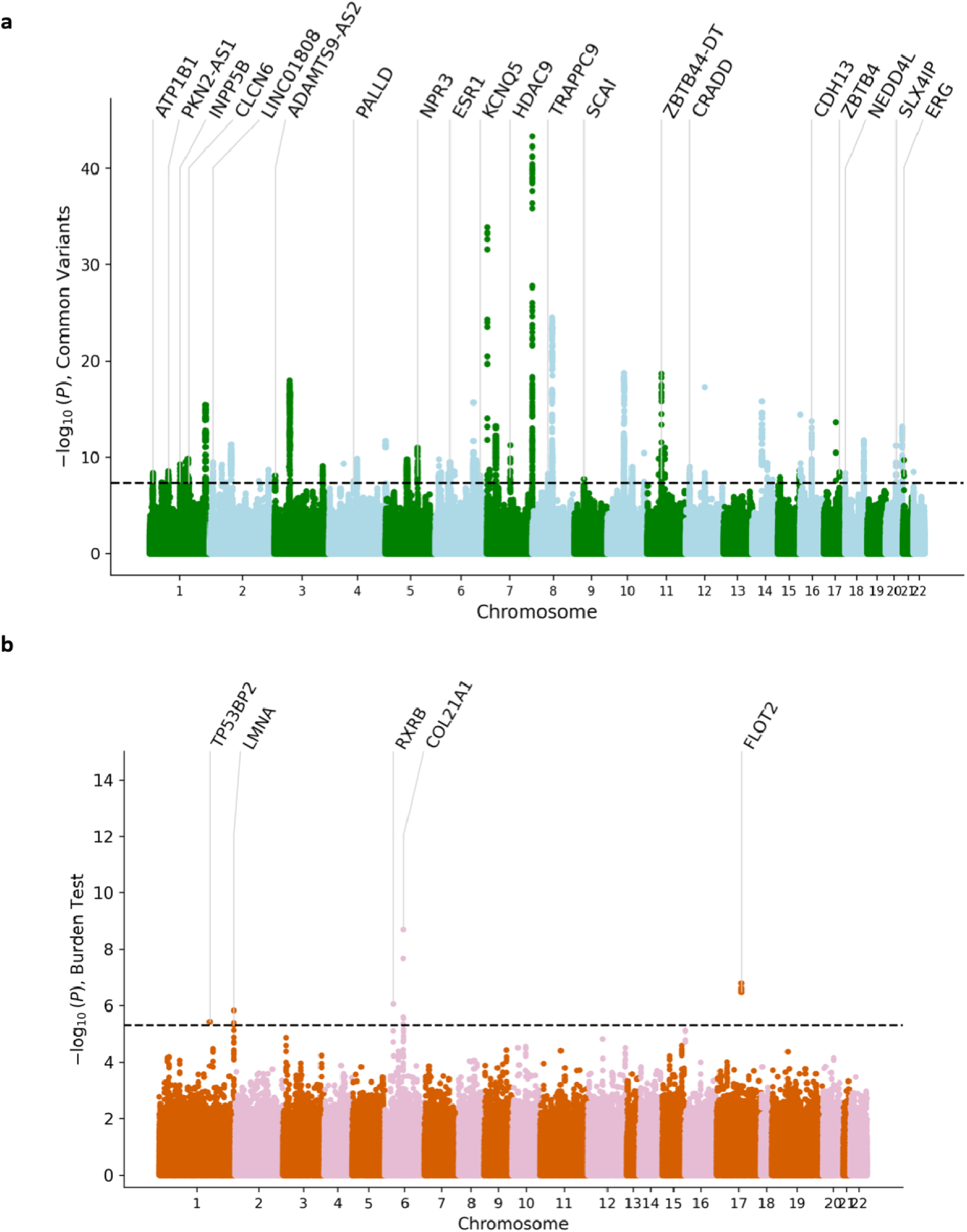
Genomic Analyses of *ArtAgeGap*. (a) Common variant genome-wide association study (GWAS) Manhattan plot (n = 114,098) displaying the -log□□(*p*) of single nucleotide polymorphism (SNP) associations with *ArtAgeGap* across autosomes 1-22. The horizontal dashed line marks the genome-wide significance threshold (*p* < 5×10□□). Peaks correspond to genome-wide significant loci after linkage disequilibrium (LD)-based clumping (r² < 0.001 within 10 Mb), with the top 20 loci annotated with the nearest genes. (b) Rare variant burden testing Manhattan plot (n = 110,326), showing -log□□(*p*) for gene-based association tests aggregating coding variants with minor allele frequency (MAF) ≤ 1% under multiple functional masks. The dotted line indicates exome-wide significance (*p* < 5×10□□).

To assess the influence of rare coding variation, we also performed gene□based burden tests in 110,326 participants with WES data by collapsing rare variants (MAF ≤ 1%, 0.1%, 0.01%) across four functional masks (predicted loss□of□function (pLoF) + missense variants with REVEL >0.5; all missense variants; pLoF only; missense variants with REVEL >0.5) and tested associations with *ArtAgeGap* (**Figure 5b**). At the exome□wide significance threshold (p < 5×10^−6^), five genes showed significant associations. The strongest association was observed for *COL21A1* under the pLoF + missense mask, where each additional variant increased *ArtAgeGap* by 0.86 years (SE=0.14; p=1.99×10□□); notably, pLoF variants in *COL21A1* have previously been also linked to BP^18^. Additional significant associations included *FLOT2* (all missense, p=1.61×10□□), *RXRB* (pLoF + missense, p=8.78×10□□), *LMNA* (all missense, p=1.46×10□□), and *TP53BP2* (pLoF only, p=3.68×10□□). *LMNA* mutations are well□established causes of cardiomyopathies^19^. These five high□confidence genes represent promising candidates for functional follow□up in arterial aging.

### Integration with human arterial transcriptomics reveals fibroblast enrichment of prioritized genes

As most lead variants in our GWAS were located within non-coding regions and our results pointed to enrichment in arterial tissue, we next integrated the GWAS data with expression quantitative trait loci (eQTL) data from human arteries (aorta and coronary arteries) in GTEx. For gene□level MR analyses, we selected genetic instruments from each gene’s GWAS summary statistics by filtering SNPs with p□<□5×10^−8^ and applying linkage disequilibrium (LD) clumping (r^2^ <□0.001 within a 10□Mb window). Using these instruments, TWAS-MR analyses in aorta (**Supplementary table S16**) identified significant associations of genetically proxied expression of 159 of 6,267 tested genes (with available instruments within the tested tissues) with *ArtAgeGap* at Benjamini-Hochberg false discovery rate (FDR)□<□0.05. Furthermore, genetically proxied expression of 58 out of 2,755 tested genes reached FDR□<□0.05 in coronary arteries (**Supplementary Table S18**). To distinguish shared causal variants from MR associations driven by linkage, we performed Bayesian colocalization between gene expression and *ArtAgeGap* at the loci of the significant genes. In aortic tissue (**Supplementary Table S17**), 83 of 159 genes showed strong evidence of colocalization (PP_4_≥0.8), indicating that the same variant likely influences both gene expression and *ArtAgeGap*. The top colocalized loci (PP_4_=1.00) included *CTB-111H14.1*, *ULK4*, *PRKAR2B*, *PHACTR1*, and *CCDC71L*. In coronary arteries (**Supplementary Table S19**), 34 of 58 genes colocalized (PP_4_≥0.8) with *ULK4*, *CTB-111H14.1*, *HSD52*, *PHACTR1*, and *FHL3* showing the strongest evidence of colocalization (all PP_4_≥0.99). Notably, 25 genes, including *INPP5B*, *ATP1B1*, *MFSD13A*, and *SF3A3*, showed significant colocalization with gene expression in both aorta and coronary arteries, strengthening their candidacy as causal mediators of the genetic signals for arterial aging.

To triangulate evidence for the involvement of the significant genes in arterial aging, we also explored, whether their expression in aorta or coronary artery changes with age. Using GTEx bulk RNA-seq data, we fitted generalized linear models to assess relationships between age at sample collection and gene expression (**Supplementary Table 20-21**). A total of 28 genes showed significant associations with age in the same direction as their genetically predicted expression was associated with *ArtAgeGap* (**Supplementary Table 22**). Among the 28 prioritized genes, *ULK4* and *RGS19* stood out, exhibiting consistent directionality across all three lines of evidence and in both tissues (**Figure 6a**). To understand which cell types, drive the expression of these genes in human arteries, we investigated open-access single-cell RNA sequencing data from human arterial samples. Fibroblasts emerged as by far the most enriched cell type for the expression of 26 of 28 significant genes. (**Figure 6b**). *RGS19* and *OPRL1* showed stronger enrichment in macrophages.

**Figure 6.**
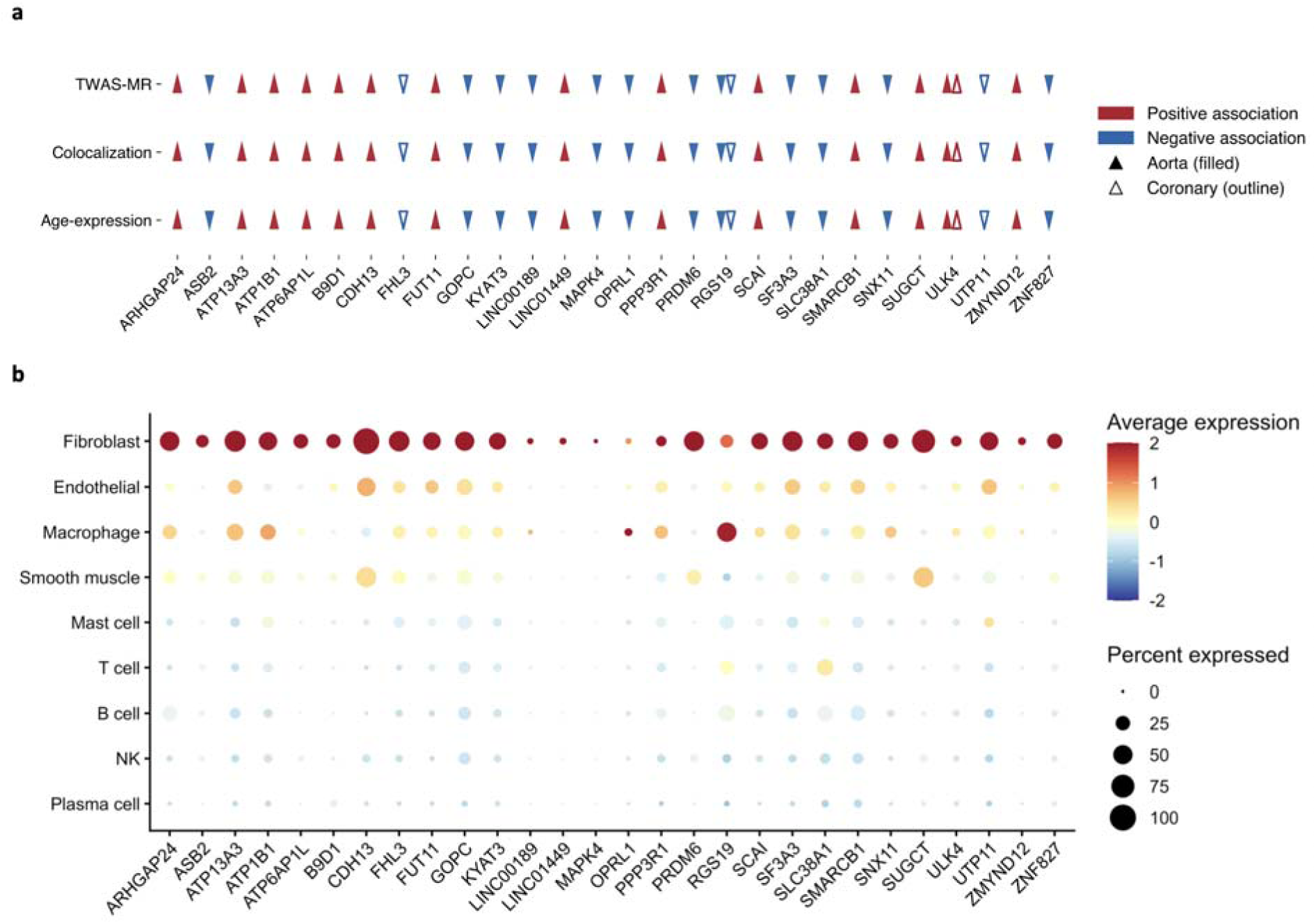
Integrative analyses of genomic and transcriptomic data in human arteries. **(a)** Summary of genes showing significant associations with arterial aging in either aortic or coronary artery tissues across three different analyses: transcriptome-wide association study-mendelian randomization (TWAS-MR) of genetically proxied expression in the GTEx resource and *ArtAgeGap*; colocalization between the gene expression and *ArtAgeGap* at the gene *cis*-locus; associations between gene expression in the respective tissue and chronological age at time of sample collection (n=472 for aorta and n= 268 for coronary arteries). **(b)** Dot plot showing the average expression (color scale) and percentage of expressing cells (dot size) of prioritized genes across major vascular and immune cell types in human carotid arteries, as derived from a meta-analysis of available single-cell RNA sequencing data.

### Mendelian randomization points to genetic associations of arterial aging with vascular risk factors and cardiovascular outcomes

We next applied MR to assess potentially causal effects of vascular risk factors on *ArtAgeGap* (**Figure 7a**, **Supplementary Table S23**). Using inverse-variance weighted (IVW) MR as the primary approach, we observed strong positive associations for central adiposity and cardiometabolic traits. Each standard deviation (SD) increased in genetically proxied waist-hip ratio was associated with a 0.59-year higher *ArtAgeGap* (P = 2.6×10□□). BP showed similarly robust effects: per 1-mmHg increase, diastolic pressure raised *ArtAgeGap* by 0.05 years (p=0.0014) and systolic pressure by 0.18 years (p=9.4×10□¹³³). Genetic liability to type 2 diabetes (yes vs. no) was also linked to higher *ArtAgeGap* (P = 1.9×10□□). For lipid traits, IVW MR indicated that a 1-SD increase in HDL cholesterol reduced *ArtAgeGap* by 0.17 years (p=0.013), whereas LDL cholesterol was not significant by IVW but suggested a positive effect in MR-Egger (+0.23 years, p=0.022). We also tested if genetically proxied *ArtAgeGap* is associated with major cardiovascular outcomes. The primary IVW MR analysis showed significant effects on multiple endpoints, including coronary artery disease (odds ratio □OR] per 1-year increment = 1.05, 95% CI: 1.01-1.10), carotid plaque (OR=1.15, 95%CI: 1.10-1.20), any stroke (OR=1.04, 95% CI: 1.01-1.07), ischemic stroke (OR=1.04, 95%CI: 1.01-1.06), large artery atherosclerotic stroke (OR=1.10, 95%CI: 1.02-1.19),heart failure (OR=1.03, 95%CI: 1.01-1.04), and peripheral artery disease (OR=1.09, 95%CI: 1.05-1.14, **Figure 7b**, **Supplementary Table S24**). Sensitivity analyses largely supported the main MR results. Although Cochran’s Q revealed some heterogeneity across traits, Egger intercept tests provided no strong evidence of directional pleiotropy, indicating that the findings were not driven by directional pleiotropy.

**Figure 7:**
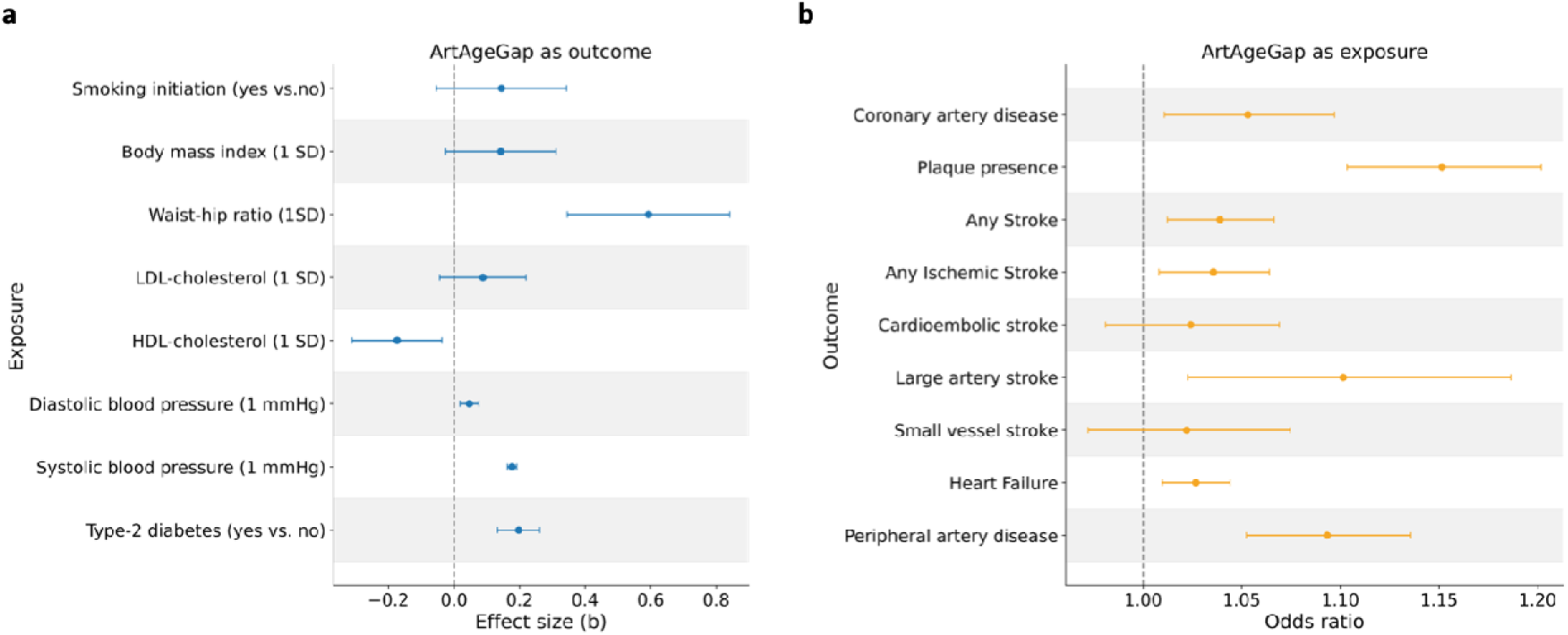
Mendelian Randomization of *ArtAgeGap* as outcome and exposure. (a) Forward mendelian randomization (MR) forest plot showing estimates of associations of genetic proxied smoking initiation, body mass index (BMI,) waist-hip ratio (WHR), LDL-cholesterol, HDL-cholesterol, systolic and diastolic blood pressure (SBP, DBP), and type 2 diabetes (T2D), with *ArtAgeGap*. Each bar represents an effect size (b) per unit increase in the genetic instrument, as indicated in the brackets, with 95% confidence intervals. (b) Forest plot of inverse-variance weighted MR associations between genetically proxied *ArtAgeGap* (per +1 year) and major cardiovascular outcomes. Odds ratios (OR) and 95% confidence intervals (CI) are displayed.

## Discussion

In this study, we introduce a deep learning-derived arterial aging clock based on PPG waveforms and BP assessments. The resulting biomarker, *ArtAgeGap*, captures inter-individual variation in arterial aging, correlates with vascular risk factors, and predicts future cardiovascular risk beyond chronological age. With a heritability of 18%, genetic analyses identified both common and rare variants associated with *ArtAgeGap,* implicating genes enriched for expression in arterial tissue. Integration with human arterial transcriptomic data further prioritized 28 potentially causal mediators of arterial aging, predominantly expressed in fibroblasts, pointing to cell-type-specific pathways as promising targets for interventions to slow vascular aging.

Developing *ArtAgeGap* within the deeply-phenotyped UK Biobank enabled the largest genomic exploration to date of an arterial aging phenotype. Among the 60 loci discovered through GWAS, several overlapped with loci previously linked to vascular traits, such as hypertension (*NPR3, SCAI, JCAD*), pulse pressure and arterial stiffness (*PKN2-AS1*, *CLCN6, TRAPPC9, CRADD, NFAT5*), aortic geometry (*SLC24A3, SCAI, LINC02398),* electrocardiography traits (*MYH6, CCDC141, SCN5A, SLC35F1*), and atherosclerotic disease (*HDAC9, JCAD*). MR analyses further supported associations between *ArtAgeGap* and both vascular risk factors and cardiovascular outcomes, validating that at the genetic level *ArtAgeGap* captures a largely arterial pathology.

More than half of the identified loci (34 out of 60) were novel and have not been previously linked to vascular pathologies. The observation that genes nearest to the top variants are most enriched for expression in arterial tissue suggests that these variants may act within the vascular wall. Extensive integrative analyses of genomic and transcriptomic data prioritized several largely unexplored candidate genes, variation in the expression of which may drive aspects of arterial aging and represent potential molecular entry points for therapeutic intervention. Among them, *ULK4*, a serine/threonine kinase involved in autophagy, harbors polymorphisms associated with blood pressure variation, and higher arterial expression correlates with delayed onset of type B aortic dissection, suggesting a protective role in maintaining vascular integrity during aging^20,21^. RGS19 belongs to the regulators of G-protein signaling (RGS) family, which modulate signaling via G-protein-coupled receptors, and its expression has been associated with coronary artery disease^22^. Knockout of *RGS19* in human hepatocytes *in vitro* led to marked reductions in the secretion of APOB100, cholesterol, and triglycerides into the culture medium, suggesting a potential role in lipid metabolism^23^. At the cellular level, single-cell RNA sequencing revealed that the vast majority of the prioritized genes were preferentially expressed in fibroblasts. As key regulators of extracellular matrix organization, fibroblasts are integral to the structural and mechanical properties of the arterial wall^24^. Given the established role of matrix remodeling and pro-fibrotic pathways in arterial stiffening^25^, this observation provides a compelling rationale for downstream experiments to elucidate how variation in these genes contributes to arterial aging.

Beyond common regulatory variation, our rare variant analyses revealed five genes (*COL21A1*, *LMNA*, *FLOT2*, *RXRB*, *TP53BP2*) whose coding variation contributes to inter-individual differences in arterial aging. Among these, *COL21A1* encodes collagen type XXI, a basement membrane-associated collagen largely localized in the arterial wall^26^. Loss-of-function or damaging missense variants in this gene increased *ArtAgeGap*, consistent with its proposed role in maintaining arterial tensile strength and integrity^27^. *LMNA*, a structural nuclear envelope protein, is mutated in laminopathies characterized by premature vascular aging and cardiac hypertrophy. *LMNA* mutational models show vascular smooth muscle cell depletion, increased medial collagen and arterial stiffening^28^. *FLOT2* is a membrane□associated scaffolding protein enriched in lipid rafts of vascular endothelial cells, where it supports flow□mediated translocation of integrin□α5 under atheroprone shear stress, thereby influencing mechanosensitive arterial signaling^29^. *RXRB*, encoding the retinoid X receptor beta, modulates transcriptional programs of lipid metabolism and inflammation^30^, key contributors to atherosclerosis and age-related vascular remodeling and stiffening. Finally, *TP53BP2*, encoding ASPP2, a regulator of p53-dependent apoptosis, has not been previously linked to vascular disease, but has been shown to anti-fibrogenic properties through inhibition of TGF-β1-induced autophagy^31^. Together, both common and rare variant analyses converge on mechanisms of extracellular matrix organization, mechanotransduction, and fibrosis regulation, reinforcing the central role of matrix-regulatory processes in arterial aging.

Our study also has implications for arterial aging phenotyping. *ArtAgeGap* estimation can be applied widely in population-based cohorts with available BP and PPG assessments, offering a scalable approach for biological discovery. Its biological relevance is highlighted by its robust associations with incident hypertension, adverse cardiovascular events, and vascular mortality. Despite being based on easily obtainable measures, *ArtAgeGap* achieved predictive performance (r = 0.60) comparable to previous arterial aging clocks derived from tonometry-based pulse waveforms^32^, MRI-derived 3D aortic geometry^33^, or artery-specific proteomic signatures^34^. Expanding PPG phenotyping from a single assessment to dynamic, serial monitoring could reduce noise and capture additional variability relevant to arterial aging. Consistent with this, recent work using 30-day PPG monitoring at the wrist via a wearable device achieved even stronger prediction of chronological age (r = 0.95).^16^ The affordability and broad availability of PPG make it particularly well-suited for large-scale studies. Whether PPG-based assessment of arterial aging could have clinical utility beyond longevity research applications should be explored in future work.

Our work has limitations. First, although the UK Biobank provides a rich resource, its specific characteristics may limit generalizability. The training sample was restricted to individuals aged 40-74 years and is largely composed of White British participants. Extending training to younger, older, and more ethnically diverse cohorts will be important in future studies. Second, the MAE of chronological age prediction (5.61) is comparable to that of other aging clocks^35,36^, but indicates room for improvement. Reliance on a single PPG waveform, which can be influenced by temporary external exposures (e.g., smoking, caffeine, sleep behavior, skin temperature), could introduce potential measurement variability in our analyses. Third, PPG and BP capture primarily functional aspects of arterial aging and may miss early structural changes in the arterial wall. Integrating accessible imaging modalities, such as vascular ultrasound to assess arterial wall thickness and composition, could further enhance arterial aging predictions.

In conclusion, we developed a PPG-based arterial aging clock that captures vascular pathology and cardiovascular risk, offering a scalable framework for population-level studies of arterial aging biology. Our genetic analyses reveal a substantial heritable component implicating previously unrecognized mechanisms, particularly those involved in fibrosis regulation and extracellular matrix organization. These findings directly inform future translational studies by highlighting candidate therapeutic targets for slowing arterial aging.

## Methods

### Study Population

The UK Biobank is a large-scale, prospective cohort study comprising approximately 501,938 individuals aged 40 to 69 years at recruitment, conducted between 2006 and 2010. Participants underwent comprehensive phenotyping, including detailed questionnaires, physical examinations, genotype characterization, biological sample collection, accelerometry, and multimodal imaging. Longitudinal follow-up for a broad array of health outcomes is available through electronic health records and death registries. PPG measurements were obtained *via* finger pulse oximetry in a subset of participants across up to four assessment visits^37^.

In total, the UK Biobank includes 254,731 PPG waveform observations from 219,453 individual participants. Alongside raw waveforms, derived characteristics and hemodynamic measures such as pulse rate and blood pressure were also collected. In this workflow, the term “observation” refers to individual waveform recordings (rather than unique participant), as we used four visits to train the model, and some participants contributed data across multiple timepoints. For validation and downstream analyses, we restricted our analyses to instance 0 (baseline visit) to ensure consistency and to avoid duplication across participants.

For model development and validation, we included observations with complete PPG waveform data, blood pressure data, and key covariates **(Figure 1b**). Participants taking anti-hypertensive medications were excluded from all stages of the analysis to avoid treatment-induced bias in PPG waveforms^38^. To ensure waveform quality, we applied the EclipseEnvelope algorithm implemented in Python to identify and remove outlier waveforms. The remaining observations were stratified by chronological age into training and testing datasets. The final model training and testing sets comprised 14,315 and 10,710 observations, respectively. Descriptive characteristics of both datasets are presented in **Supplementary Tables S1 and S2**. For downstream clinical and genetic analyses, we retained all eligible observations from instance 0, excluding those used for model training and those with extreme predicted arterial ages (defined as <30 or >80 years), yielding a final dataset of 114,344 observations. Genetic analyses were conducted on a subset of 114,098 individuals with available genotyping data. DNA was extracted from peripheral blood and genotyped using a custom Affymetrix UK Biobank Axiom array, capturing 812,428 biallelic SNPs and insertions/deletions (indels). Genotype imputation was performed using a combined reference panel from the Haplotype Reference Consortium (HRC), the UK10K, and the 1000 Genomes Project, resulting in a final dataset of over 90 million variants available for association and downstream analyses.

### Photoplethysmography Pulse Waveform Acquisition

PPG pulse waveforms were recorded using the PulseTrace PCA2 device (CareFusion, USA), which employs an infrared sensor attached to the participant’s finger. The resulting waveform captures dynamic fluctuations in peripheral blood volume, representing both the forward-traveling arterial pulse wave originating from the upper-body arterial system and its subsequent reflection from the lower-body vasculature. Each recording session lasted approximately 10-15 seconds. Participants were seated in an upright position with their feet flat and parallel on the floor, and toes pointing forward. The left arm was fully supported and rested in a relaxed, supinated position on the desktop surface. The sensor clip was placed on a finger of the left hand. To promote autonomic relaxation and minimize signal variability, participants were instructed to take five deep breaths before measurement initiation. Waveform acquisition began once a stable signal was observed on the device monitor, ensuring optimal data quality. Further details regarding the acquisition protocol are reported in the UK Biobank Resource 100240 (available at https://biobank.ndph.ox.ac.uk/ukb/ukb/docs/Pulsewave.pdf).

### Data Processing

Two primary data modalities were utilized for the deep learning-based model development: (1) PPG pulse waveforms and (2) tabular hemodynamic and demographic features. PPG waveform data were stored as strings containing 100 numerical points delimited by vertical bars (“|”). During preprocessing, these strings were parsed, the separators removed, and the points converted into 1×100 numerical vectors suitable for integration into the tabular dataset.

The detailed preprocessing steps were as follows:

#### Blood Pressure Harmonization and Pulse Pressure Calculation

Systolic and diastolic blood pressure were obtained using both automated and manual devices (UK Biobank fields 4080, 93, 4079, and 94). When both readings were available, an average across all values was computed. If only manual measurements were present, their average was prioritized; if only automated readings were available, these were used. Pulse pressure was calculated as the difference between systolic and diastolic blood pressure.

#### Precise Chronological Age Calculation

Chronological age was computed by subtracting the date of birth (field 33) from the date of assessment (field 53) and was used for both the training of the deep learning model development and calculation of the *ArtAgeGap*.

#### Exclusion of Participants on Antihypertensive Therapy

Participants prescribed antihypertensive medications were identified using medication codes (field 20003) and excluded from all datasets to eliminate treatment-induced confounding of hemodynamic parameters.

#### Exclusion of Participants with Incomplete Data

Participants with missing values for any key variables for training and analysis, whether model inputs or features used in downstream analyses were removed.

#### CVD History Annotation

Self-reported presence of CVD at baseline was assessed using the UK Biobank field 6150 (“Self-reported vascular/heart problems diagnosed by a doctor”), defined as angina, myocardial infarction, stroke, or hypertension. Participants who reported any defined diseases were labeled as 1 (‘CVD history’), while those selecting “None of the above” were labeled as 0 (‘No CVD history’). People who reported ‘Prefer not to answer’ were labeled as -3 and not included in analysis.

#### Pulse Waveform Quality Control

Waveform quality was evaluated by peak count. Approximately 2% of waveforms exhibited six or more peaks, indicative of noise or artifacts. These were excluded using the EclipseEnvelope method implemented in Python, with the threshold set at 2%.

#### Calibration to Pulse Rate

Since each PPG waveform represents a single cardiac cycle and due to variability in cardiac cycle duration across individuals, calibration was performed using pulse rate. The time gap between the waveform points was calculated as:

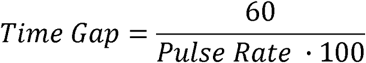

Time-dependent waveform features (e.g., notch time, peak time, shoulder time) were computed by multiplying the time gap by their respective positional indices. Calibrated waveforms were used to generate time-aligned images.

#### Stratified Sampling for Deep Learning-Based Model Development

The training dataset (n=14,315) was created by randomly selecting an equal number of observations for each chronological age across all available instances to ensure a uniform age distribution. The testing dataset (n=10,710) was similarly stratified using only instance 0 data.

#### Normalization

All continuous tabular variables were normalized to a [0, 1] range using MinMaxScaler from scikit-learn (v1.3.0). Normalization parameters were computed from the training dataset and consistently applied to the test and analysis sets.

#### Exclusion of Implausible Arterial Age Predictions

After model training, to reduce the impact of biologically implausible predictions, observations with model-PAs below 30 or above 80 years (<0.1%) were excluded from downstream analyses.

#### Software and Computational Tools

All preprocessing tasks were conducted in Python 3.11.5 using the following libraries: NumPy (v1.24.3), Pandas (v2.0.3), and scikit-learn (v1.3.0). A custom adaptation of EllipticEnvelope was used for waveform quality control.

### Multimodal Neural Network Architecture

A multimodal neural network was implemented using the PyTorch Lightning framework (v1.6.0), comprising two parallel processing branches, one for pulse waveform data and the other for tabular features.

The first branch was a CNN designed to extract embeddings from PPG waveforms. It included three convolutional blocks with progressively increasing feature dimensions (16, 32, and 64), each followed by ReLU activation. Batch normalization and dropout (dropout rate = 0.5) were applied to enhance model generalization. The resulting feature maps were flattened and passed through a fully connected layer (16, 20) with ReLU activation.

The second branch was an MLP that processes tabular hemodynamic and socio-demographic data, consisting of three fully connected layers with dimensions (111,64), (64,30), and (30,20), each followed by a ReLU activation function. The outputs from the CNN and MLP branches (each a 1×20 vector) are concatenated into a 1×40 feature vector. A ReLU activation function is applied to the concatenated vector, which is then passed through a final fully connected layer (40,1) to generate a prediction of CA.

The model was trained using the Adam optimizer (β_1_=0.9, β_2_=0.999) with a learning rate of 1×10^−3^. A batch size of 32 was used for both training and validation. The training was conducted for a maximum of 20 epochs, with early stopping applied based on validation MAE to prevent overfitting. A validation set comprising 20% of the training data was held out during training. Model performance was monitored on the validation set, and a learning rate scheduler reduced the learning rate by a factor of 0.1 if the validation MAE did not improve over 7 consecutive epochs.

For comparison and to assess the specific contribution of PPG-derived information to age prediction, we trained a gradient boosting regression model (XGBoost) to predict chronological age from only three hemodynamic features: systolic blood pressure, diastolic blood pressure, and pulse pressure. The model employed 500 estimators, a learning rate of 0.05, maximum depth of 3, subsample ratio of 0.8, and column subsampling ratio of 0.8, using five-fold cross-validation. Training and testing were performed on the same datasets as the multimodal framework. Model performance was evaluated using the coefficient of determination (R²), MAE, RMSE, averaged across folds. All analyses were conducted in Python (v3.11) using *xgboost* (v3.0.2) and *scikit-learn* (v1.5.1).

### Biomarker Generation

Initial analysis revealed a systematic bias in the model’s predictions of arterial age (PA): participants with higher chronological age (CA) tended to receive lower PA estimates, while younger individuals exhibited overestimated PAs. As a result, the unadjusted age gap (PA - CA) was negatively correlated with CA, indicating a dependency on age that could confound its interpretation as a biomarker of vascular aging.

To correct for this bias, we modeled the expected age gap across the CA distribution using a second-order polynomial regression, by calculating a standardized age gap (STA) as a function of CA as follows:

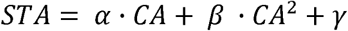

This estimated STA was then subtracted from the raw age difference to derive the final bias-corrected biomarker, *ArtAgeGap*:

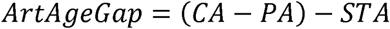

This adjustment ensured that *ArtAgeGap* was independent of CA, thereby enhancing its interpretability as a measure of individual vascular aging deviation relative to age-matched individuals.

### Assessment of Disease Status in Relation to Age Outputs

To evaluate how PA and *ArtAgeGap* varied by disease status and CA group, we visualized their distributions using violin plots stratified by CVD status and CA. Participants were grouped into six 5-year CA bins. Within each bin, separate violin plots were generated for diseased and non-diseased individuals, displaying the distribution and mean of PA and *ArtAgeGap*. Plots were generated using the matplotlib library (v3.8.4) in Python. We used a two□sided Mann-Whitney U test to assess whether the “No CVD history” and “CVD history” groups derive from the same distribution. Statistical significance is indicated by asterisks: p < 0.05 (*), p < 0.01 (**), and p < 0.001 (***). All tests were performed using the SciPy package (v1.13.1).

### Correlation with Arterial Aging Factors

We assessed monotonic associations between the CA bias□corrected age gap (*ArtAgeGap*) and a panel of hemodynamic and anthropometric measures using Spearman’s rank correlation in Python 3.10. From our dataset, we extracted *ArtAgeGap* and eleven numeric features (pulse□wave notch time, peak time, shoulder time, arterial stiffness index, diastolic and systolic blood pressure, pulse pressure, peak□to□peak interval, peak count, pulse rate, and standing height), and dropped any rows with missing values on a per□pair basis. For each feature, we computed Spearman’s ρ and a two□sided p□value via scipy.stats.spearmanr. Correlation coefficients and p□values were tabulated, and significance levels were annotated as p < 0.05 (*), p < 0.01 (**), and p < 0.001 (***).

### Association with Cardiovascular Risk Factors

To assess the clinical relevance of *ArtAgeGap*, we conducted multivariable linear regression analyses relating *ArtAgeGap* to six traditional cardiovascular risk factors. Binary variables such as statin use, smoking status (ever vs. never), and diabetes medication use were coded as 0 (no) and 1 (yes). BMI was scaled in standard deviation units (4.4 kg/m^2^), while both HDL and LDL cholesterol were scaled per 1 mmol/dL. Models were fit using ordinary least squares (OLS) via the LinearRegression class in scikit-learn (v1.3.0, Python 3.10). Regression coefficients, 95% confidence intervals (CI), and two-sided p-values were reported.

### Association with Future Vascular Events

To evaluate the predictive value of *ArtAgeGap* as a biomarker, we employed Cox proportional hazards models to estimate its association with incident vascular outcomes. Primary endpoints included AMI, stroke, heart failure, unstable angina, cardiovascular-related death, and all-cause mortality. We also analyzed a composite endpoint of these outcomes (MACE). Furthermore, we examined incident hypertension, as recorded in participants’ health records as a separate primary endpoint. Diagnoses and mortality were ascertained via ICD-9 and ICD-10 codes (**Supplementary Table 25**). Analyses were restricted to individuals free of the respective endpoint diagnosis at baseline. Time-to-event was computed from the assessment date (instance 0, “Date of Attending Assessment Centre”) to the event date (diagnosis or death). For censored cases, follow-up was calculated up to the latest update of death records (October 31, 2022). After exclusion of individuals with missing covariates, the final cohort for this analysis comprised 96,615 individuals. In a sensitivity analysis for the hypertension endpoint, to reduce misclassification due to undiagnosed cases in health records. we included only participants with repeated systolic and diastolic BP measurements across all four UK Biobank assessment visits (instances 0-3) and no evidence of hypertension at baseline. Hypertension at each visit was defined as systolic blood pressure >140 mmHg or diastolic blood pressure >90 mmHg.

Cox models included the following covariates: sex (0=female, 1=male), CA (per year), *ArtAgeGap* (per year), BMI (per kg/m^2^), statin use (0=no use, 1=use), diabetes medication use (0=no use, 1=use), Townsend Deprivation Index (per unit), previous smoke (0=no, 1=yes), current smoke (0=no, yes=1). Analyses were conducted using the lifelines package (v0.28.0) in Python. Hazard ratios (HRs) and 95% CIs were reported for CA and *ArtAgeGap*, with additional covariate results available in the Supplementary Tables.

### Common Variant Genome-Wide Association Study

We conducted a GWAS to examine the relationship between approximately one million genetic variants and the *ArtAgeGap* phenotype in 114,098 individuals with available whole-genome sequencing data. The analysis was performed using Regenie (v3.3)^39^, a two-step linear mixed model approach. In the first step, a whole-genome prediction model was constructed by partitioning genetic markers into blocks to efficiently manage memory usage. In the second step, the association between each SNP and *ArtAgeGap* was tested. All available SNPs were included in the analysis without prior filtering. The model was adjusted for sex and the first ten principal components (PCs) of genetic ancestry. While processing the results, we use a filter with MAF >1% and imputation quality (INFO score) >0.3. Heritability estimation was conducted by LDSC (v1.0.1) python package, with reference as European population^40,41,42^.

### Rare Variant Association and Gene-Based Burden Testing

To extend the genetic analysis to low-frequency and rare variants from WES data, we utilized Regenie Step 2 to perform single-variant and gene-based burden tests^39^. Rare variants were stratified by MAF (MAF ≤ 0.01, ≤ 0.001, ≤ 0.0001) and analyzed under the same covariate-adjusted linear model framework as common variants. For gene-level tests, rare alleles were collapsed into burden scores using four functional masks: (i) predicted loss-of-function (pLoF) + missense variants with REVEL > 0.5; (ii) all missense variants; (iii) pLoF variants only; and (iv) missense variants with REVEL > 0.5. Burden scores were regressed on *ArtAgeGap* using the same covariates utilized in Regenie Step 1, including sex and the first 10 PCs. Genes achieving p < 5×10^−6^ were considered significantly associated with *ArtAgeGap*.

### Linkage Disequilibrium Clumping, Gene Annotation and Novel loci Discovery

To identify independent association signals, we performed LD-based clumping using the ld_clump function from the TwoSampleMR R package. Clumping was performed using the European population reference panel from the 1000 Genomes Project, with parameters: clumping window =10 Mb (clump_kb=10,000), LD threshold=R^2^ <0.001. Within each LD block, the SNP with the smallest p-value was retained as the lead SNP.

Gene annotation for these lead SNPs was performed using ncbi_snp_query, which maps each variant to its nearest gene and provides functional annotations based on the dbSNP database, including transcript context and predicted consequence.

The clumped significant loci were input to GWAS catalog^43^, if there is no previous associations with BP, arterial stiffness and other cardiometabolic traits, they were considered as novel loci. The number of novel loci was reported.

### Mendelian Randomization

We applied two-sample MR using the TwoSampleMR R package (v0.5.6, R 4.1.0) to infer potential causal relationships. SNPs associated with exposures at genome-wide significance (p <5×10^−8^) were clumped (R² <0.001, 10 Mb window) using the 1000 Genomes European reference panel. The inverse-variance weighted (IVW) method was used for primary causal estimates, with MR-Egger and weighted median estimators as sensitivity analyses. Instrument heterogeneity and directional pleiotropy were assessed using Cochran’s Q (mr_heterogeneity) and MR-Egger intercept (mr_pleiotropy_test), respectively. When *ArtAgeGap* was treated as the outcome, we evaluated risk factors such as smoking initiation^44^, BMI^45^, waist-hip ratio^45^, LDL and HDL cholesterol^46^, BP (systolic and diastolic)^47^. When *ArtAgeGap* was the exposure, we examined cardiovascular endpoints including coronary artery disease^48^, plaque presence^49^, any stroke, and ischemic stroke subtypes (cardioembolic, large-artery, or small-vessel)^50^, periphery artery disease^51^, and heart failure^52^. (**Supplementary Table S26**).

### Transcriptome-Wide Association Study Mendelian Randomization

We performed TWAS-MR using GTEx v8 cis-eQTL data from aorta and coronary artery tissues. For each gene, we selected SNPs within ±1 Mb of the transcription start site with eQTL p <5×10^−8^. Clumping (R^2^ <0.001, 10 Mb window) was performed using the 1000 Genomes European reference. The remaining SNPs served as instruments in two-sample MR analyses testing the effect of gene expression on *ArtAgeGap*. Analyses were conducted using TwoSampleMR in R 4.1.0. IVW was the primary estimator, with MR-Egger and weighted median estimators used for sensitivity analysis. P-values were corrected for multiple testing using the FDR, and genes with FDR-adjusted values smaller than 0.05 were reported.

### Colocalization Analysis

We performed Bayesian colocalization analysis using the coloc R package to assess whether cis-eQTL signals in coronary artery tissue (GTEx v8) and GWAS signals for *ArtAgeGap* (N=114,098) were driven by a shared causal variant. For each gene with significant MR evidence (FDR q <0.05), we compiled two SNP-level summary datasets: (i) cis-eQTL statistics from GTEx, including the effect and alternate alleles, effect size (β), variance of the effect (SE^2^), MAF, and p-value; and (ii) matched GWAS summary statistics for *ArtAgeGap* with the same fields. We removed duplicate SNP entries and explicitly specified sample sizes (n=213 for cis-eQTLs; n=114,098 for GWAS). Colocalization was performed using the coloc.abf function with prior probabilities set to p_1_=p_2_=1×10^−4^ and p_12_=1×10^−5^. Posterior probabilities for hypotheses H_0_ through H_4_ were estimated, and the posterior probability of a shared causal variant (PP_4_ >0.8) was used as the primary metric of colocalization.

### Age-associated gene expression analysis

Age-related transcriptional changes were assessed using a generalized linear modeling framework in DESeq2 (v1.46.0). Raw RNA-seq counts were filtered to remove lowly expressed genes, retaining genes with (i) total counts ≥10 across all samples and (ii) non-zero counts in ≥10 samples, and normalized for library size. Gene-wise counts were modeled with a negative binomial distribution, including age as a continuous predictor and relevant covariates (e.g., sex, batch) to control for confounding. Age was represented using the midpoint of each GTEx age interval (e.g., the interval 10-20 was assigned an age of 15). For each gene, the age effect was tested with a Wald test on the age coefficient, and log□ fold changes per unit of age were stabilized using apeglm (v1.28.0) shrinkage. Multiple testing was controlled by the Benjamini-Hochberg false discovery rate; we report FDR-adjusted P values and age-associated log□ fold changes. All analyses were performed in R (DESeq2, apeglm), accessed from Python via rpy2 (v3.6.4) in a Jupyter notebook.

### Tissue Enrichment and Cell Type Enrichment

To investigate the tissue enrichment of the closest genes of clumped significant loci, we input the gene lists to FUMA (https://fuma.ctglab.nl) and chose GENE2FUNC method with GTEx v8^53^. For cell type-specific enrichment, we utilized the Single Cell Portal^54^ (https://singlecell.zendesk.com/hc/en-us) and identified two relevant single-cell RNA sequencing datasets: one is carotid artery tissue^55^, and another is aorta tissue^56^. The top-ranked genes from our multilayer prioritization analysis were queried to determine in which cell types they were significantly more highly expressed compared with other cell types (log□(FC) ≥ 0.26, adjusted P < 0.05).

To investigate cell type enrichment of prioritized genes, four published human carotid artery single-cell RNA-seq datasets were reprocessed using a uniform Seurat (v4.3) workflow^57,58,59,60^. Cells with fewer than 300 detected genes, fewer than 1,000 total counts, or greater than 20% mitochondrial reads were excluded. Data were normalized with SCTransform, doublets were removed using DoubletFinder (expected doublet rate 5%), and datasets were integrated with Harmony on the top 10 principal components, accounting for sample and sequencing platform as covariates. Clustering was performed using the Leiden algorithm (resolution = 1.3). Cell-type identities were assigned by manual marker-based annotation using the level-1 cell-type definitions and marker lists described previously^61^. Cluster-level normalized expression values from the SCTransform assay were used to determine cell-type-specific gene expression.

## Data Availability

Individual-level data and genome data from UK Biobank (application number :36993, 7089, and 151281) are available under controlled access to protect participant confidentiality and in line with consent agreements. Approved researchers can apply for access through the UK Biobank data access framework (https://www.ukbiobank.ac.uk/enable-your-research/apply-for-access). The full dataset is hosted on the UK Biobank Research Analysis Platform (https://www.ukbiobank.ac.uk/enable-your-research/research-analysis-platform). GWAS summary statistics for *ArtAgeGap* will be made publicly available through the GWAS Catalog.

## Code Availability

The code used to build the model in this study will be made publicly available on GitHub upon publication of this manuscript.

